# Implementing a clinical pathway for diagnosing and treating acute HIV infection among key populations attending sexual health clinics in Indonesia: cohort profile of the INTERACT study

**DOI:** 10.1101/2024.06.06.24307250

**Authors:** Irwanto, Nurhayati H. Kawi, Hendry Luis, Erik P. Sihotang, Pande Putu Januraga, Margareta Oktaviani, Suwarti, Dwi P. Rahmawati, Evi Sukmaningrum, Evy Yanihastuti, Maartje Dijkstra, Eduard J. Sanders, F. Stephen Wignall, Keerti Gedela, Raph L. Hamers, INTERACT Study Group

**Author notes:** KG and RLH are shared last authors. Members listed at end of paper (INTERACT Study Group). **Correspondence to** Raph L Hamers, MD PhD, Oxford University Clinical Research Unit Indonesia, Faculty of Medicine Universitas Indonesia, Jalan Salemba Raya No. 6, 10430 Jakarta Pusat, Indonesia.

## Abstract

**Background:** To reduce the high HIV incidence among key populations in Indonesia, we implemented a clinical pathway for screening, diagnosis and treatment of acute HIV infection (AHI) in sexual health clinics in Jakarta and Bali. This paper presents a cohort profile and analysis of baseline data on the study uptake, diagnostic yield, and estimated AHI prevalence and screening cascade outcomes.

**Methods:** We performed a baseline analysis of 1879 individuals who underwent AHI screening at three sexual health clinics in Jakarta and Bali between May and December 2023, comprising a risk-score assessment, fourth-generation antibody/p24 antigen-based rapid diagnostic test (RDT; Abbott Determine HIV Early Detect) and HIV-PCR (Xpert) testing.

**Results:** Median age was 27 years (IQR24-31), and 75.4% were male. Men who have sex with men (MSM) accounted for 50.4%, clients of sex workers 20.1%, and sex workers 5.2%. Of 1866 participants tested at study enrolment, 113 (6.1% [113/1866]) had chronic HIV (antibody-positive) and 6 (0.34% [6/1748]) had AEHI. HIV-PCR testing led to a 5.3% (95%CI1.9-11.2) increase in confirmed HIV diagnoses. The number needed to test to detect one AEHI case was 291 (1748/6) overall and 169 (842/5) among MSM. Overall HIV and AHI prevalence was 6.4% (95%CI 5.3-7.6; 119/1866) and 0.34% (95%CI0.12-0.74; 6/1748) overall; and 10.8% (95%CI8.9-13.0; 102/940) and 0.53% (95%CI 0.17-1.2; 5/940) among MSM. The Abbott Determine HIV Early Detect RDT only detected 2 (18.2%) of 11 AEHI cases. 113 (95.0%) newly diagnosed individuals were linked to care and started ART, of whom 75 (66.4%) on the same day and 104 (92.0%) within a week (median 0 days, range 0-93).

**Conclusion:** AHI screening, diagnosis and prompt treatment is feasible among high-risk urban MSM in Indonesia. Further evaluations are needed to estimate clinical impact and cost-effectiveness of AHI screening in this setting. The study continues accrual and follow-up, and provides a platform for future immuno-virological, social science, and intervention studies in Indonesia.

## INTRODUCTION

### Background

Indonesia is a socio-culturally, economically and geographically diverse, Muslim-majority, middle-income country with the world’s fourth largest population (275 million), featuring stark health inequalities across regions and communities. Indonesia has one of the highest numbers of new HIV infections globally, estimated at 24,000 (95% confidence interval [CI] 22,000-27,000) in 2022 (1), with an estimated 515,455 persons living with HIV (PLHIV) by March 2023 (2). Although recent estimates have shown a decreasing trend overall, numbers of new HIV infections have remained high among vulnerable groups, specifically men who have sex with men (MSM), transgender women (TGW), and female sex workers and their sexual partners. The most recent national survey data (2018-2019) estimated a national HIV prevalence of 17.9% (95%CI 16.8-19.0) for MSM and 11.9% (95%CI 10.7-13.1) for TGW (3), with province-level estimates for MSM of 23.0% in Jakarta and 21.2% in Bali (3). A retrospective analysis among MSM and TGW found a very high HIV incidence of 9.4 per 100 person-years (95%CI7.9-11.2) in Jakarta and 7.2 per 100 person-years (95%CI5.7-9.1) in Bali (4). Access to oral HIV pre-exposure prophylaxis (PrEP), currently being rolled out in many other countries (5), remains largely restricted to a pilot programme in a limited number of government primary health centres (2).

In Indonesia, HIV care cascades for testing, diagnosing, linkage to care and suppressive treatment are often fractured, particularly for key populations (6,7). By March 2023, national data estimated that, overall, 85% of PLHIV knew their HIV status, 36% received antiretroviral therapy (ART), and 10% had viral load suppression on ART (7). In an earlier study in 2015-2016 among 637 MSM diagnosed with HIV attending non-government sexual health clinics, 83% were linked to care, 73% started ART, 57% were retained in care, 43% had a viral load test done after six months, and 39% had viral load suppression on ART (6). There are numerous social, economic and structural factors underlying the fractured HIV care cascades (8,9). The government prioritises integration of HIV services in general primary health centres, where high levels of stigma and discrimination against MSM, TGW and PLHIV have been reported, whereas sexual health services tailored to key populations are only offered by a few private and non-government clinics. Social stigmatization, discrimination, economic deprivation and punitive laws further increase the vulnerability to HIV and hinder public health access in these settings (8,9).

### Acute HIV infection as a driver of epidemics

Acute HIV infection (AHI) is the short phase immediately after viral acquisition, and is typically characterised by a lack of anti-HIV antibodies and the presence of viremia, which can be detected by an HIV-RNA or p24 viral antigen test (10). However, clinicians often fail to recognize AHI because most people experience no or non-specific symptoms. The rate of sexual transmission has been estimated to be 8-20 times higher during AHI than during chronic infection (11–13), and phylogenetic and mathematical models, mainly from Europe and the Americas, have estimated that AHI may account for 10-50% of all new HIV infections among MSM communities (14,15). Reliable estimates for AHI as a driver of the HIV epidemic among Asian key populations are lacking, although a modelling study among MSM in Bangkok estimated that AHI detection and immediate ART initiation could reduce onward transmissions by 89% (16). To reach the UNAIDS goals towards HIV elimination (17), it is imperative that all PLHIV are diagnosed early and start ART immediately, to prevent onward transmission (18,19). Undiagnosed AHI may hinder achieving the desired population benefits associated with those comprehensive “Treat all” strategies (14).

### Study rationale

#### AHI-centred combined intervention models

From a public health perspective, AHI screening is an important strategy to decrease HIV transmission via viral load reduction and behavioural interventions, provided that individuals with AHI can be diagnosed, linked to care and started on same-day or immediate ART (16,17); to improve uptake of pre-exposure prophylaxis among HIV-negative persons at high risk of HIV acquisition (20,21); and to enhance partner services via notification of persons recently exposed or likely transmitting (14,22,23). Moreover, from a clinical perspective, there are substantial immunological and virological benefits to identifying and treating persons with AHI, evading irreversible damage to host immune systems (24,25) and seeding of viral reservoirs that occurs during untreated AHI (14,25,26).

Experiences in urban MSM communities in, for instance, San Francisco (27), British Columbia (28), London (29) and Amsterdam (30), indicate that combined prevention and treatment interventions (including for AHI) tailored to key populations with city-specific strategies to remove structural barriers to access services are most successful in reducing incident HIV infections and containing HIV epidemics. These experiences create a strong impetus to tailor such successful models to populations with ongoing HIV transmission in low- and middle-income countries (LMIC).

#### Sensitive AHI screening approaches

Third-generation HIV antibody rapid diagnostic tests are the backbone of testing in Indonesia, as in many LMICs, missing the earliest pre-antibody phase that is AHI. Fourth-generation HIV-antigen/antibody combination tests and nucleic acid (HIV-PCR) tests shorten the post-transmission detection window from 2-6 weeks (antibody), to 15-20 days (antigen) or even 7-10 days (HIV-PCR). However, although fourth-generation antigen/antibody point-of care assays are more straightforward and cheaper than HIV-PCR, they have shown variable sensitivities for detecting AHI (28-88% across cohorts), (31–33). Optimal AHI algorithms must balance the consequences of missed diagnoses and cost, speed and ease-of-use. Many AHI testing algorithms combine third and/or fourth generation assays with more sensitive HIV-PCR testing technologies. Risk score algorithms based on symptoms and/or sexual risk behaviour have been developed to optimize the efficiency and reduce cost of AHI screening approaches, but have been mainly validated on retrospective datasets from Western and African MSM communities. The diagnostic yield of targeted HIV-PCR testing has been estimated at 3.3% (95%CI 2.2-4.6%) across three studies (34). Further prospective studies are needed to assess and optimize the yield of AHI risk score and testing algorithms in a variety of settings and populations. Pooled (group) sample testing has been successfully applied to increase testing efficiency and reduce required resources (31,35,36).

### Study objectives

The aim of INTERACT is to assess whether implementing an AHI test and treat clinical pathway at sexual health clinics serving Indonesian MSM and other key populations, coupled with a digital community engagement tool tailored to the wider target community, can strengthen the HIV care cascade in Jakarta and Bali. Specific objectives are:

1. To assess the effectiveness/yield as well as the uptake, acceptability and feasibility, and barriers and enablers of the AHI test and treat clinical pathway;
2. To assess whether uptake of the intervention can be enhanced through a tailored digital engagement intervention;
3. To estimate the potential population impact on HIV incidence and cost-effectiveness of the AHI test and treat clinical pathway in Indonesia.

## METHODS

### Design

INTERACT is a prospective cohort study of individuals aged 16 years or older who are vulnerable to HIV acquisition and are voluntarily attending an open-access sexual health service in Jakarta or Bali for HIV testing. INTERACT is a collaboration between researchers and care providers involving several Indonesian and international institutions with track records in HIV prevention and care models. Study management is done by the Oxford University Clinical Research Unit (OUCRU) Indonesia, based in Jakarta. Ethics approvals were obtained from the Atma Jaya Catholic University research ethics committee (0009R/III/PPPE.PM.10.05/10/2022) and the Oxford Tropical Research Ethics Committee (565-22). The study enrolment period is from May 2023 through December 2024. This study is reported as per Strengthening the Reporting of Observational Studies in Epidemiology guidelines (**Table S1**).

The central guiding principle for study implementation is that INTERACT procedures are integrated into the routine client services, compliant with the Ministry of Health HIV guidelines. The INTERACT AHI test and treat clinical pathway intervention comprises the following (**Figure 1**):

- Implementation of an AHI screening approach. A participant-completed AHI risk assessment (“Risk Checker”), modified from the Amsterdam AHI risk score (37), is followed by Xpert HIV-PCR diagnostic testing, with same-visit or next-day delivery of results. Participants are encouraged to return for regular (three-monthly or earlier in case of AHI risk exposure and/or symptoms) AHI screen visits;
- Offer of immediate, same-visit ART initiation in newly HIV diagnosed persons, and monitoring HIV-RNA at 3 and 6 months after ART initiation to assess adherence and virological response;
- Offer of assisted partner notification, to identify and test sexual partners at high risk of undiagnosed HIV;
- Implementation of a tailored digital engagement intervention, through social media platforms, which aims to promote uptake of AHI testing at the study clinics (www.CekUpYuk.id, promoted since March 2024).

**Figure 1.**
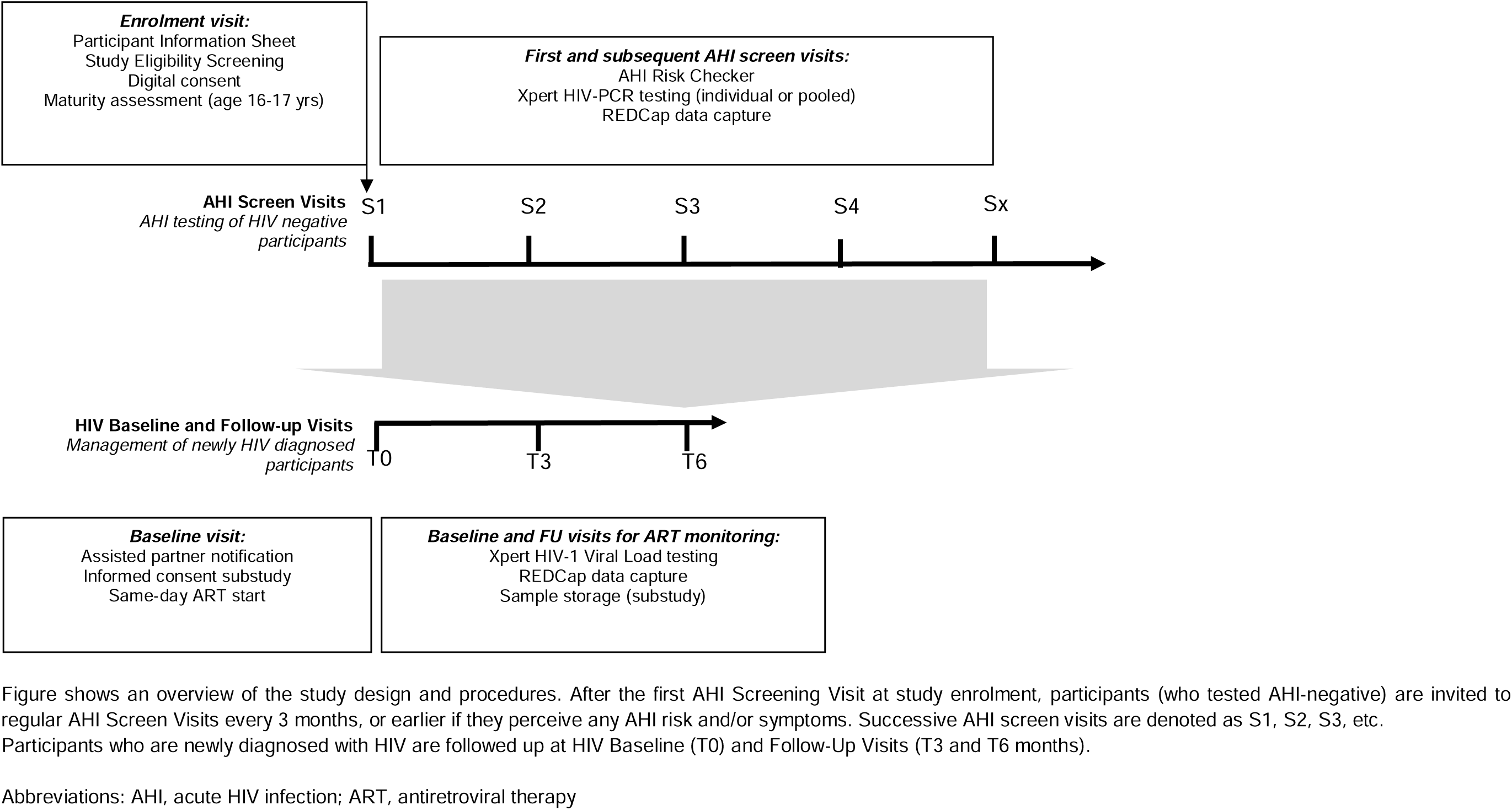
Study design overview

Several inter-related quantitative and qualitative outcomes will be measured, related to the effectiveness/yield, uptake, acceptability, feasibility and potential impact of the AHI clinical pathway (**Table S2)**. An implementation evaluation, based on stakeholder interviews (38), will help identify barriers and enablers. The study data will be used to perform a cost-effectiveness analysis, estimate the role of AHI as a driver of the local epidemic, and predict the impact of scalable intervention scenarios on local epidemic. A subset of newly HIV diagnosed individuals will be enrolled, with written informed consent, in a viro-immunologic longitudinal sub-study to investigate AHI immune responses, antiretroviral drug resistance patterns and HIV transmission networks.

### Setting and population

INTERACT is implemented at three open-access NGO sexual health clinics providing free or low-cost HIV and other STI testing and treatment services, predominantly serving MSM, TGW, and sex workers and their clients. Yayasan Bali Peduli (www.balipeduli.org), operates clinics in Denpasar and Ubud, Bali, and Klinik Utama Globalindo (www.yayasankasihglobalindo.org) operates in South Jakarta. DKI Jakarta and Bali are among the provinces with the highest reported HIV prevalence (**Figure S1**). All sites offer point-of-care rapid antigen/antibody HIV testing (Abbott Determine HIV Early Detect) and HIV-PCR testing (Xpert HIV Viral Load and HIV Qual). Assuming that ∼50% of the HIV-antibody-negative participants classify as AHI high-risk (N∼2250) at 3.3% (95%CI 2.2-4.6) yield for targeted HIV-PCR testing (34), we expect to detect 75 (95%CI 50-104) AHI cases at an absolute precision of ±4%, given alpha 5%. Assuming that the remaining ∼50% of the HIV-antibody-negative participants classify as non AHI high-risk (N∼2250) at a 0.2% (95%CI 0.1-0.3) yield for universal HIV-PCR testing (34), we expect to detect an additional 5 (95%CI 2-7) AHI cases through pooled-sample screening.

### Study screening and eligibility assessment

All individuals who attend the clinic for HIV testing, both first-time and returning, are informed about the ongoing INTERACT project through a participant information sheet and study banner in the waiting room. Interested individuals are invited to answer study eligibility questions and provide digital consent into a REDCap electronic data capture on a clinic-provided or personal mobile device. Eligibility criteria are:

- Aged 16 years or older (individuals aged 16 or 17 years undergo an additional maturity assessment); AND
- Individuals not known to be living with HIV; AND
- Individual belongs to one or more of the following HIV risk categories:

◦ Men who have sex with men
◦ Transgender women
◦ Persons who inject drugs
◦ Sex workers
◦ Clients of sex workers
◦ Sexual partners of PLHIV
◦ Other risk of HIV acquisition (undisclosed category)

Eligible, consenting individuals are consecutively enrolled and proceed to AHI screening procedures. For individuals who are not eligible or decline participation, the reason is recorded. All clinic attendees receive services as usual as per standard clinic procedures.

### AHI screen visit procedures

At each AHI screen visit, participants are asked to fill out the “AHI Risk Checker”, modified from the Amsterdam AHI risk score (37): three or more sexual partners, any STI, condomless receptive anal sex (each within the past six months); weight loss, fever, swollen lymph nodes, oral thrush (each in the past two weeks). After completion, the client responses are verified between the client and a counsellor. An AHI risk score is automatically calculated by summing up the number of risk factors and/or symptoms (range 0-7). Individual HIV-PCR testing is applied to participants classified as “AHI high-risk” based on their risk score (the initial cutoff >=1 was adjusted to >=2 in October 2024, to strive for ∼50% being classified as “AHI high-risk”), whereas all other participants are HIV-PCR tested using a pooled sampling approach (see below).

Specimens that test HIV-antigen/antibody-positive or indeterminate (i.e. Abbott Determine HIV Early Detect or equivalent) undergo confirmatory testing with a third-generation HIV-antibody RDT (i.e. Bioline HIV1/2, or equivalent). The study applies additional HIV-PCR testing to all specimens that are HIV antigen and antibody negative or inconclusive/indeterminate, using a study-specific SOP (**Figure S2**). Participants classified as AHI high-risk receive an Xpert HIV-1 Qual test on an individual sample; if positive confirmed with Xpert HIV-1 Viral Load assay (final result reported on the same day, or <24h). Participants not classified as AHI high-risk receive an Xpert HIV-1 Viral Load on a pooled sample; if positive, deconvolution testing of the constituent individual samples with Xpert HIV-1 Viral Load assay; if positive, confirmed with Xpert HIV-1 Qual assay (final result reported <72h). All positive HIV-PCR results are confirmed on a new specimen. Confirmed HIV diagnoses are classified as either acute (AHI) (PCR and/or p24 positive, antibody-negative, including antibody-discordance or early HIV), recent HIV (antibody-positive and documented negative test <6 months prior), or chronic HIV (antibody-positive and no documented negative test <6 months prior) (**Table S3**). Participants who test HIV-negative are invited (with text message reminders) to regular follow-up AHI screen visits (every three months or earlier if they perceive any AHI risk or symptoms). **Table S4** summarises the clinical an lab variables being collected.

### Follow-up visit procedures for participants newly diagnosed with HIV

Participants who are newly diagnosed with HIV (including AHI) receive follow-up visits, according to existing clinic procedures (**Figure 1**). During the baseline visit (T0), same-day ART initiation and assisted partner notification are offered. The standard first-line ART regimen is emtricitabine 200mg/tenofovir disoproxil fumarate 245 mg/dolutegravir 50 mg [TLD] once daily. Xpert HIV Viral Load is measured before ART start, and at 3 and 6 months after ART start. Participants in the immuno-virologic sub-study provide a blood sample for storage.

### Patient and public involvement

The design of the AHI clinical pathway involved clinic staff who are part of or interact closely with the key populations they serve, including PLHIV. A community advisory board, consisting of representatives of PLHIV and MSM community organisations, has been established to help inform and design community engagement tools, share study findings and identify further research priorities. Knowledge dissemination strategies prioritise sharing findings with health policymakers and communities and fostering a dialogue that may guide research direction and further enhance collaboration.

## FINDINGS TO DATE

### Study uptake and enrolment

This paper summarises the baseline data of participants enrolled in the first eight months of the study. From May 12 to December 31, 2023, of overall 5262 individuals seeking HIV testing across the sites, 1946 (37.0%) were screened for study eligibility (1621 of 3789 [42.8%] in Jakarta and 325 of 1473 [22.1%] in Bali), whereas 3022 refused (most without providing a reason) and 294 were not offered study eligibility screening (mainly because research staff were not available or the client came outside of laboratory service hours) (**Figure 2**). Of 1946 individuals screened for study eligibility, nearly all (1879, 96.6%) were eligible and enrolled (1559 in Jakarta and 320 in Bali); the reasons for not enrolling were previously tested HIV-positive (41), not willing to provide consent (12), and not reporting any HIV risk (6).

**Figure 2.**
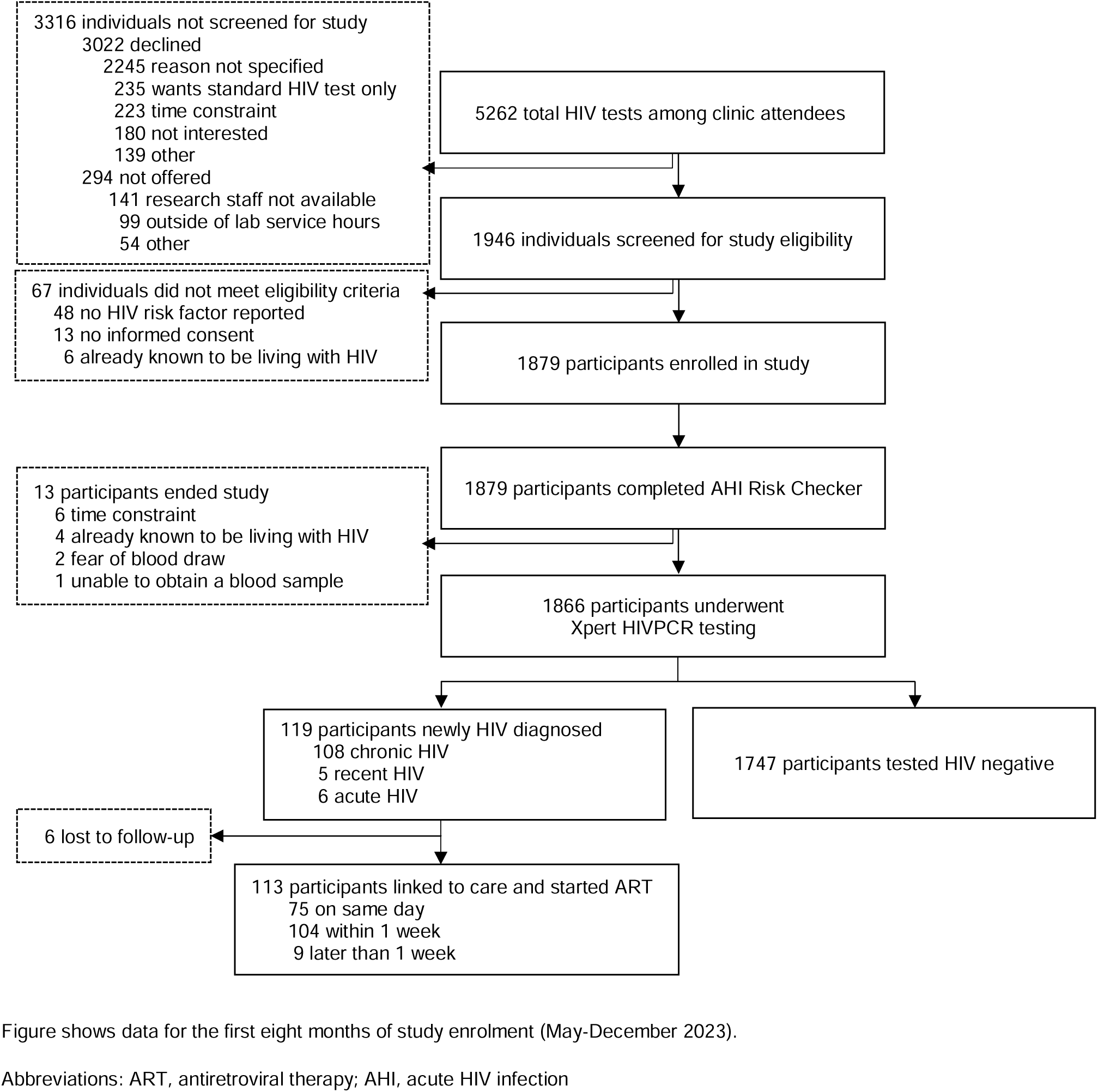
Study flow for the first AHI screen visit (study enrolment)

**Figure 3.**
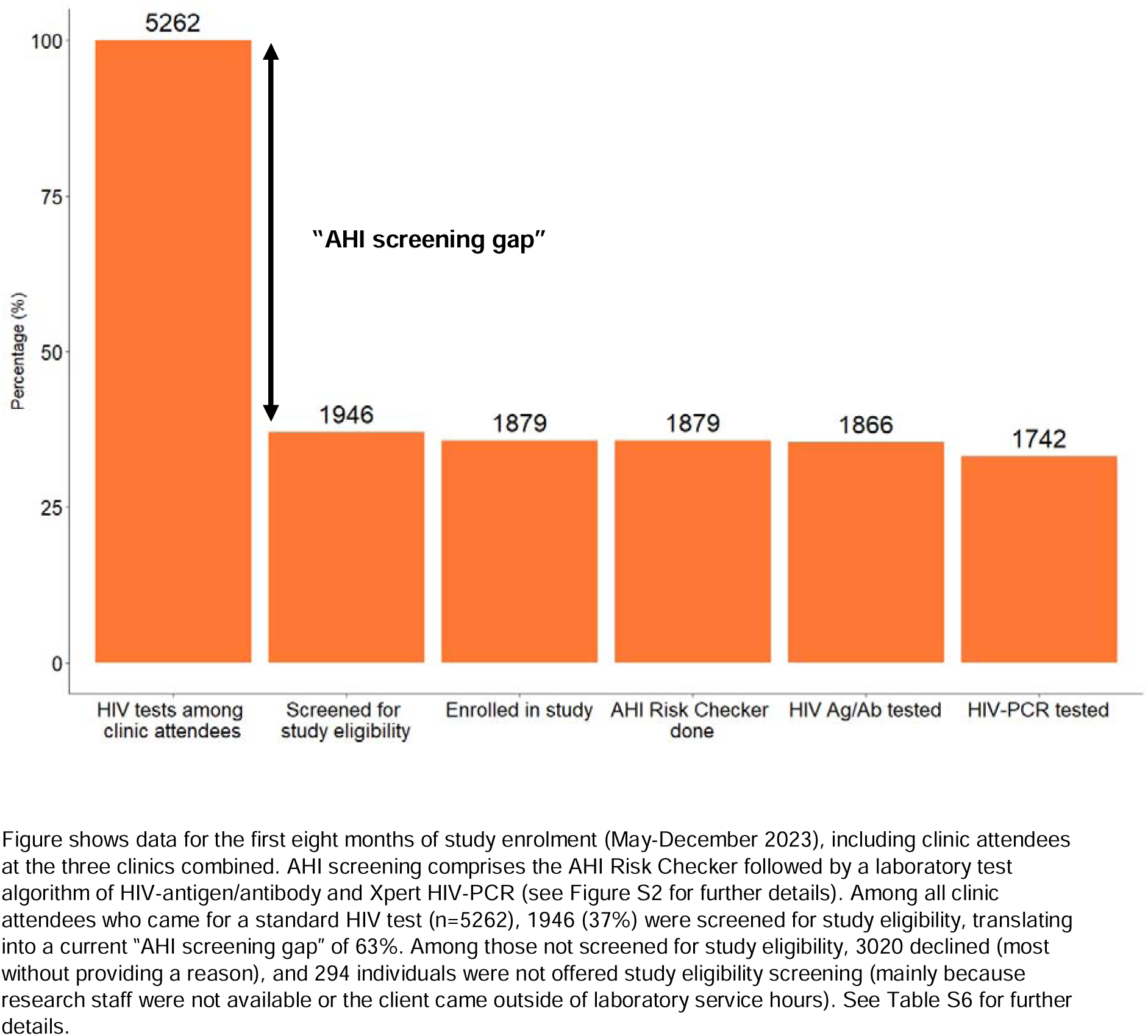
AHI screening cascade

### Participant characteristics at enrolment

**Table 1** summarises the characteristics of the 1879 participants. 1416 (75.4%) were male, 439 (23.4%) female, 15 (0.8%) transgender women, and 9 (0.5%) identified as other gender. The median age was 27 years (IQR 24-31). Most participants completed university/higher education (1335, 71.0%) and were employed (1588, 84.5%). The most reported HIV risk categories was MSM (947, 50.4%), followed by sex worker client (378, 20.1%), having a sexual partner living with HIV (143, 7.6%), sex worker (97, 5.2%), transgender woman (15, 0.8%), person who inject drugs (6, 0.3%), and those not disclosing their risk category (296, 15.8%). 1003 (53.4%) participants had a previous negative HIV test result, of whom nearly half (444, 44.3%) within the past six months. Main reasons for current HIV testing included feeling at risk (1156, 61.5%), having symptoms (456, 24.3%), retesting because of window period after possible recent exposure (261, 13.9%), a new sexual relationship (252, 13.4%), getting married (132, 7.0%), or having a partner who tested HIV positive (77, 4.1%).

**Table 1.** Participant characteristics at enrolment.

| Variable | Individuals screened for study eligibility<br>N=1946 | Participants enrolled in the study<br>N=1879 | P-value<sup>1</sup> | Participants who tested HIV-negative<br>N=1747 | Participants who tested HIV-positive<br>N=119 | P-value<sup>2</sup> |
| --- | --- | --- | --- | --- | --- | --- |
| **Age (years)** |  |  |  |  |  |  |
| Median (IQR) | 27 (24-31) | 27 (24-31) | 0.900 | 27 (24-31) | 27 (24-30) | >0.999 |
| 16-19 | 89 (4.6%) | 87 (4.6%) | 0.999 | 573 (32.8%) | 5 (4.2%) | 0.942 |
| 20-24 | 635 (32.6%) | 617 (32.8%) |  | 506 (28.9%) | 41 (34.5%) |  |
| 25-29 | 573 (29.4%) | 548 (29.2%) |  | 369 (21.2%) | 37 (31.1%) |  |
| 30-34 | 407 (20.9%) | 395 (21%) |  | 217 (12.4%) | 22 (18.5%) |  |
| ≥35 | 242 (12.4%) | 232 (12.3%) |  | 82 (4.7%) | 14 (11.8%) |  |
| **Sex at birth** |  |  |  |  |  |  |
| Male | 1509 (77.5%) | 1457 (77.5%) | >0.999 | 1331 (76.2%) | 115 (96.6%) | <0.001 |
| Female | 437 (22.5%) | 422 (22.5%) |  | 416 (23.8%) | 4 (3.4%) |  |
| **Gender identity<sup>3</sup>** |  |  |  |  |  |  |
| Male | 1416 (75.4%) | 1416 (75.4%) | <0.001 | 1294 (74.0%) | 111 (93.4%) | <0.001 |
| Female | 439 (23.4%) | 439 (23.4%) |  | 431 (24.7%) | 6 (5.0%) |  |
| Transgender | 15 (0.8%) | 15 (0.8%) |  | 14 (0.8%) | 1 (0.8%) |  |
| Other<sup>4</sup> | 9 (0.5%) | 9 (0.5%) |  | 8 (0.5%) | 1 (0.8%) |  |
| **Clinic visit status** |  |  |  |  |  |  |
| First-time | 966 (49.6%) | 925 (49.2%) | 0.824 | 834 (47.8%) | 85 (71.4%) | <0.001 |
| Returning | 980 (50.4%) | 954 (50.8%) |  | 913 (52.2%) | 34 (28.6%) |  |
| **Education level** |  |  |  |  |  |  |
| Higher education/University | 1381 (71.0%) | 1335 (71.0%) | 0.962 | 1251 (71.6%) | 76 (63.9%) | 0.091 |
| High school completed | 507 (26.0%) | 487 (25.9%) |  | 449 (25.7%) | 36 (30.3%) |  |
| Middle school completed | 41 (2.1%) | 41 (2.3%) |  | 35 (2.0%) | 6 (5.0%) |  |
| Primary school completed | 6 (0.3%) | 6 (0.3%) |  | 5 (0.3%) | 1 (0.8%) |  |
| Primary school incomplete | 1 (0.1%) | 0 (0.0%) |  | 0 (0.0%) | 0 (0.0%) |  |
| Not provided | 10 (0.5%) | 10 (0.5%) |  | 7 (0.4%) | 0 (0.0%) |  |
| **Current occupation** |  |  |  |  |  |  |
| Employed | 1645 (84.5%) | 1588 (84.5%) | 0.999 | 1487 (85.1%) | 92 (77.3%) | 0.077 |
| Student | 228 (11.7%) | 220 (11.7%) |  | 201 (11.5%) | 19 (16.0%) |  |
| Unemployed | 57 (3.0%) | 55 (2.9%) |  | 47 (2.7%) | 7 (5.9%) |  |
| Unspecified | 16 (0.8%) | 16 (0.9%) |  | 12 (0.7%) | 1 (0.8%) |  |
| **Location** |  |  |  |  |  |  |
| Jakarta | 1621 (83.3%) | 1559 (83.0%) | 0.958 | 1455 (83.3%) | 96 (80.7%) | 0.339 |
| Denpasar, Bali | 248 (12.7%) | 243 (12.9%) |  | 223 (12.7%) | 15 (12.6%) |  |
| Ubud, Bali | 77 (4.0%) | 77 (4.1%) |  | 69 (4.0%) | 8 (6.7%) |  |
| **Key population<sup>5</sup>** |  |  |  |  |  |  |
| Men who have sex with men | 959 (49.3%) | 947 (50.4%) | <0.001 | 838 (48%) | 102 (85.7%) | <0.001 |
| Sex worker clients | 382 (19.6%) | 378 (20.1%) |  | 361 (20.7%) | 15 (12.6%) |  |
| Sexual partner living with HIV | 144 (7.4%) | 143 (7.6%) |  | 131 (7.5%) | 12 (10.1%) |  |
| Sex workers | 98 (5.0%) | 97 (5.2%) |  | 90 (5.2%) | 5 (4.2%) |  |
| Transgender women | 15 (0.8%) | 15 (0.8%) |  | 14 (0.8%) | 1 (0.8%) |  |
| Persons who inject drugs | 6 (0.3%) | 6 (0.3%) |  | 6 (0.3%) | 0 (0.0%) |  |
| Undisclosed risk category | 296 (15.2%) | 296 (15.8%) |  | 291 (16.7%) | 4 (3.4%) |  |
| **Previous negative HIV test** |  |  |  |  |  |  |
| Ever | 1003 (51.5%) | 1003 (53.4%) | <0.001 | 970 (55.5%) | 32 (26.9%) | <0.001 |
| In the past 6 months | 444 (44.3%) | 444 (44.3%) |  | 438 (45.2%) | 6 (18.8%) |  |
| **Reason for current HIV testing<sup>5</sup>** |  |  |  |  |  |  |
| Feeling at risk | - | 1156 (61.5%) | <0.001 | 1056 (60.7%) | 85 (71.4%) | <0.001 |
| Having symptoms | - | 456 (24.3%) |  | 433 (24.9%) | 22 (18.5%) |  |
| Retest (window period) | - | 261 (13.9%) |  | 226 (13.0%) | 32 (26.9%) |  |
| New sexual relationship | - | 252 (13.4%) |  | 246 (14.1%) | 6 (5.0%) |  |
| Unspecified | - | 225 (3.6%) |  | 216 (12.4%) | 6 (5.0%) |  |
| Getting married | - | 132 (7.0%) |  | 126 (7.2%) | 5 (4.2%) |  |
| Partner tested HIV positive | - | 77 (4.1%) |  | 68 (3.9%) | 9 (7.6%) |  |
| Partner has STI | - | 29 (1.5%) |  | 26 (1.5%) | 3 (2.5%) |  |
| Pregnant or partner pregnant | - | 7 (0.4%) |  | 7 (0.4%) | 0 (0.0%) |  |

Table shows baseline characteristics of those screened and enrolled at Globalindo Jakarta (May 12-December 31, 2023); Bali Peduli Ubud (May 30-December 31, 2023); and Bali Peduli Denpasar (May 31-December 31, 2023)
<sup>1</sup> Comparison of individuals screened versus enrolled (Chi<sup>2</sup>, Kruskal Wallis-H test, and Dunn's pairwise comparison)
<sup>2</sup> Comparison of participants who tested HIV negative versus positive (Chi<sup>2</sup>, Kruskal Wallis-H test and Dunn's pairwise comparison)
<sup>3</sup> Gender identity was not recorded for ineligible individuals
<sup>4</sup> Includes individuals who identified as non-binary or gender-fluid
<sup>5</sup> Individuals could choose more than one category

**Table 2** summarises behavioural risk factors for HIV acquisition among the study population. Around half of the participants (884, 47.0%) reported to have engaged in anal sex in the past three months, comprising receptive (306, 34.6%), insertive (316, 35.8%) or both (262, 29.6%). Condomless receptive anal sex was reported by about half of MSM (52.8%, 500/947), 5.2% (49/932) of men who did not identify as MSM, and 10.1% (36/356) of individuals not disclosing their risk category. The use of recreational drugs before or during sex (“chemsex”) in the past three months was reported infrequently (33, 1.8%), mostly “poppers” (inhaled alkyl nitrites). Engaging in group sex and visiting sex parties was reported by 106 (5.6%) and 41 (2.2%), respectively. PrEP use was reported by 156 (8.3%) participants, evenly divided between daily (76, 48.7%) and event-driven dosing (80, 51.3%), and 63 (40.4%) had last used PrEP more than a month ago. Most participants (113, 72.4%) sourced their PrEP through a government primary healthcare centre.

**Table 2.** Sexual practices and other AHI risk factors among participants at enrolment.

| Variable | All study participants<br>N=1879 | Participants who tested HIV-negative<br>N=1747<sup>1</sup> | Participants who tested HIV-positive<br>N=119<sup>2</sup> | P value<sup>3</sup> | Participants who tested AHI-positive<br>N=6 | P value<sup>4</sup> |
| --- | --- | --- | --- | --- | --- | --- |
| **AHI risk score assessment<sup>5</sup>** |  |  |  |  |  |  |
| AHI risk score (median, IQR)) | 1 (1-2) | 1(1-2) | 2(1-3) | 0.323 | 2 (1-3) | <0.001 |
| Three or more sexual partners in the past 6 months | 906 (48.2%) | 844 (48.3%) | 58 (48.7%) | 0.866 | 2 (33.3%) | 0.688 |
| Condomless receptive anal sex in the past 6 months | 585 (31.1%) | 502 (28.7%) | 77 (64.7%) | <0.001 | 4 (66.7%) | 0.062 |
| STI and/or STI symptoms in the past 6 months | 443 (23.6%) | 418 (23.9%) | 22 (18.5%) | 0.173 | 1 (16.7%) | >0.999 |
| Fever in the past 2 weeks | 391 (20.8%) | 336 (19.2%) | 52 (43.7%) | <0.001 | 4 (66.7%) | 0.015 |
| Oral thrush in the past 2 weeks | 212 (11.3%) | 192 (11.0%) | 20 (16.8%) | 0.113 | 1 (16.7%) | 0.504 |
| Weight loss in the past 2 weeks | 92 (4.9%) | 69 (3.9%) | 23 (19.3%) | <0.001 | 1 (16.7%) | 0.217 |
| Lymph nodes in the past 2 weeks | 91 (4.8%) | 76 (4.4%) | 14 (11.8%) | <0.001 | 0 (0.0%) | >0.999 |
| **Number of sex partners in the past 6 months (median, IQR)** | 2 (1-5) | 2 (1-5) | 2 (1-5) | >0.999 | 2 (2-4) | >0.999 |
| **Anal sex in the past 3 months** | 884 (47.0%) | 784 (44.9%) | 93 (78.2%) | <0.001 | 6 (100.0%) | 0.022 |
| Receptive/bottom | 306 (34.6%) | 262 (33.4%) | 41 (44.1%) | <0.001 | 3 (50.0%) | 0.009 |
| Insertive/top | 316 (35.8%) | 299 (38.2%) | 16 (17.2%) |  | 2 (33.3%) |  |
| Both insertive/top and receptive/bottom | 262 (29.6%) | 223 (28.4%) | 36 (38.7%) |  | 1 (16.7%) |  |
| **Ever injected drug use** | 6 (0.3%) | 6 (0.3%) | 0 (0.0%) | 0.690 | 0 (0.0%) | >0.999 |
| **Recreational drug use before or during sex in the past 3 months<sup>6</sup>** | 33 (1.8%) | 28 (1.6%) | 5 (4.2%) | 0.079 | 1 (16.7%) | 0.095 |
| **Group sex in the past 3 months** | 106 (5.6%) | 98 (5.6%) | 8 (6.7%) | 0.758 | 0 (0.0%) | >0.999 |
| **Visited sex party in the past 3 months** | 41 (2.2%) | 40 (2.3%) | 1 (0.8%) | 0.344 | 0 (0.0%) | >0.999 |
| **PrEP** |  |  |  |  |  |  |
| Ever used PrEP<sup>7</sup> | 156 (8.3%) | 150 (8.6%) | 5 (4.2%) | 0.209 | 1 (100.0%) | 0.418 |
| More than a month ago | 93 (59.6%) | 89 (59.3%) | 4 (80.0%) |  | 1 (100.0%) |  |
| Less than a month ago | 63 (40.4%) | 61 (40.7%) | 1 (20.0%) |  | 0 (0.0%) |  |
| PrEP regimen used |  |  |  |  |  |  |
| Event-driven dosing | 80 (51.3%) | 70 (46.7%) | 3 (60.0%) | 0.161 | 1 (100.0%) | 0.243 |
| Daily dosing | 76 (48.7%) | 80 (53.3%) | 2 (40.0%) |  | 0 (0.0%) |  |

Table summarizes the distribution of behavioural risk factors for HIV acquisition in the study population, stratified by those who tested HIV negative versus positive (including AHI), and those who tested HIV positive versus AHI positive
<sup>1</sup> 13 of 1879 (0.7%) participants did not receive an HIV-antigen/antibody test
<sup>2</sup> Comprises 113 individuals with chronic HIV (antibody positive) and 6 individuals with acute HIV (Xpert HIV-PCR-positive, antibody-negative)
<sup>3</sup> Comparison of participants who tested HIV negative versus positive (Chi<sup>2</sup> and Kruskal Wallis-H)
<sup>4</sup> Comparison of participants who tested HIV-negative versus AHI-positive (Chi<sup>2</sup> or Fischer exact and Kruskal Wallis-H)
<sup>5</sup> Modified Amsterdam AHI risk score (reference 37)
<sup>6</sup> Types of drugs used included poppers (30, 90.9%), GHB and cannabis/marijuana (each 3, 9.1%), crystal meth (2, 6.1%), benzodiazepines and ecstasy/MDMA (each 1, 3.0%)
<sup>7</sup> Mode of PrEP access included primary health centre (113, 72.4%), private clinic (29, 18.6%), ordered online (5, 3.2%), hospital (3, 1.9%), other (6, 3.8%)
Abbreviations: AHI, acute HIV infection; PrEP, pre-exposure prophylaxis for HIV

### AHI Risk Checker and laboratory test results

Figure 3 summarises the AHI screening cascade. All 1879 participants completed the AHI Risk Checker, within a median of 6.3 minutes (IQR5.0-8.4). The median AHI risk score was 1 (IQR1-2; range 0-7), comprising 906 (48.2%) reporting three or more sex partners, 585 (31.1%) condomless receptive anal sex, 443 (23.6%) STI or STI symptoms (each in the past six months), 391 (20.8%) fever, 212 (11.3%) oral thrush, 92 (4.9%) weight loss, and 91 (4.8%) enlarged lymph nodes (**Table 2**).

Of 1879 participants, 1866 (99.3%) underwent an HIV-antigen/antibody screen test. Of 1866 participants tested, 113 (6.1% [113/1866]) had recent/chronic HIV infection (antibody-positive) and 1753 who tested negative or inconclusive. Of those, 1748 (99.7%) underwent an Xpert HIV-PCR test, of whom 6 (0.34% [6/1748]) had acute HIV (Xpert HIV-PCR-positive, antibody-negative; none were antibody-discordant or early HIV). The number needed to test to detect one case was 17 (1866/113) for recent/chronic HIV infection and 291 (1748/6) for AHI overall, and, among MSM, 10 (940/97) and 169 (842/5), respectively. Xpert HIV-PCR testing led to a 5.3% (95%CI 1.9-11.2; 119/113) increase in confirmed HIV diagnoses overall, and, among MSM, 5.2% (95%CI 1.7-11.6; 102/97). HIV prevalence was 6.4% (95%CI 5.3-7.6; 119/1866) overall and, among MSM, 10.8% (95%CI 8.9-13.0; 102/940), and 6.2% (95%CI 5.0-7.5; 96/1551) in Jakarta and 7.3% (95%CI 4.7-10.7; 23/315) in Bali. AHI prevalence was 0.34% (95%CI 0.12-0.74; 6/1748) overall, and 0.53% (95%CI 0.17-1.2; 5/940) among MSM.

All six participants with AHI were below 30 years of age, five were MSM and one did not disclose their HIV risk category (**Table S5**). The median AHI risk score was 2 (IQR 1-3; range 0-6), which was statistically significantly higher than those who tested HIV-negative (1, IQR1-2; range 0-7; p<0.001), and comprised 4 (66.7%) individuals reporting condomless receptive anal sex, 4 (66.7%) fever, 2 (33.3%) three or more sexual partners, and 1 (16.7%) each an STI history, oral thrush or weight loss (**Table 2**). Assisted partner notification was only accepted by one participant with AHI, who notified three sexual partners.

### Linkage to care and ART initiation

The total time from starting AHI screening to receiving the test results was a median of 2.0 hours (IQR 1.2-2.9) for HIV-antigen/antibody testing, 2.9 (IQR 2.5-3.7) hours for individual Xpert HIV-PCR, and 5.4 hours (IQR 3.4-8.5) for pooled Xpert HIV-PCR (p<0.001). Overall, 90.1% (1590/1748) of participants received their Xpert HIV-PCR result on the same day, 96.2% (1681/1748) within 24 hours, and 99.3% (1736/1748) within 72 hours (Figure 4).

**Figure 4.**
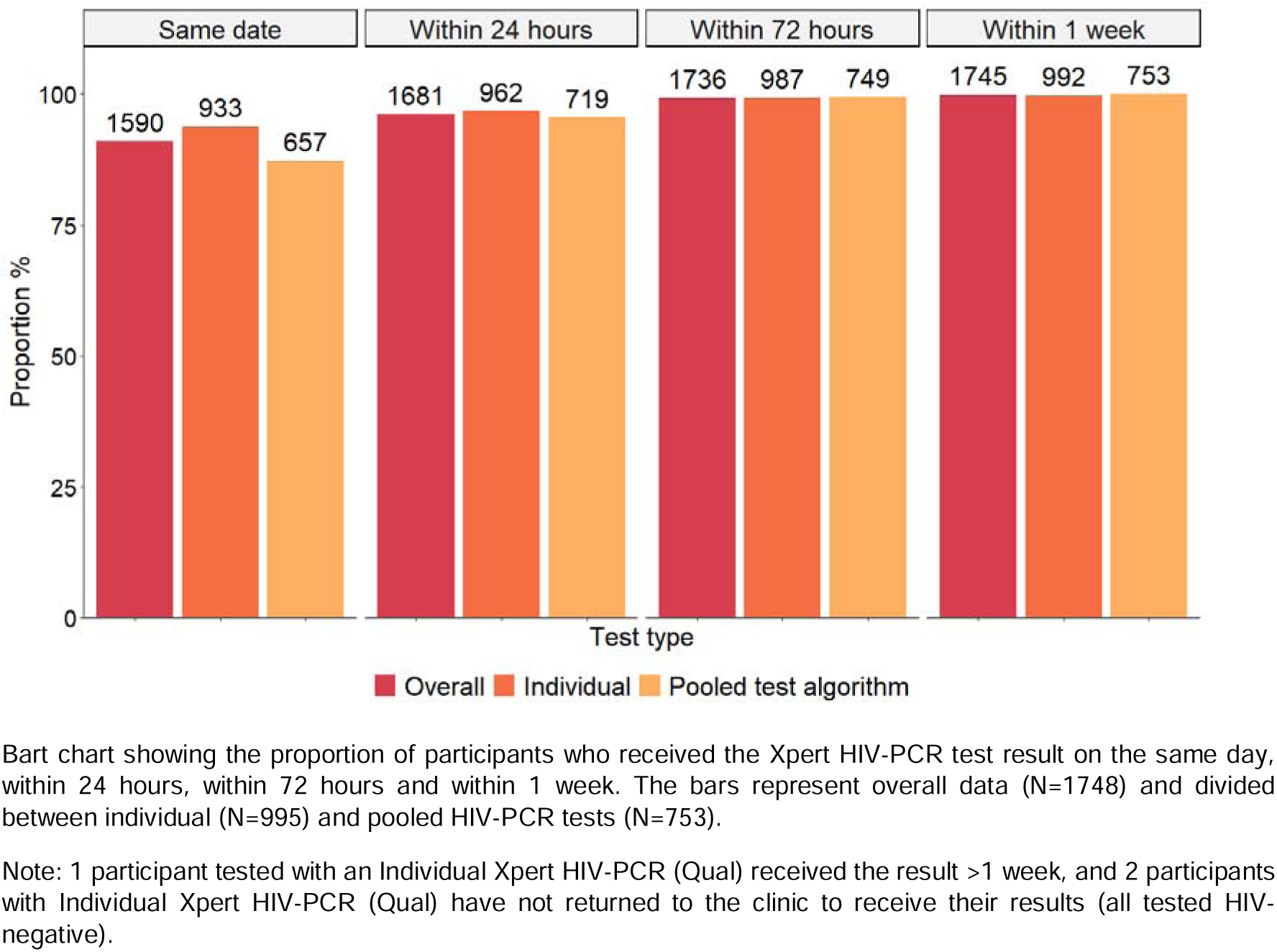
Time to receive HIV-PCR test result

Of the 119 individuals newly diagnosed with HIV (including 6 with AHI), 113 (95.0%) were linked to care and started ART, of whom 92 in the same clinic and 21 in another clinic; 75 (66.4%) started ART on the same day and 104 (92.0%) within one week (median 0 days, range 0-93), whereas 9 (8.0%) deferred ART because of a concurrent opportunistic infection (3) or referral elsewhere (6) (Figure 2). Six of the newly HIV-diagnosed individuals (including 3 with AHI) did not return to the clinic and were lost to follow-up.

## CONCLUSIONS

This cross-sectional analysis among MSM and other key populations attending sexual health clinics in Bali and Jakarta, Indonesia, has demonstrated that implementing an AHI risk score assessment and HIV-PCR-based AHI screening was feasible, and that AHI was highly prevalent among young urban MSM at 0.53% (95%CI 0.17-1.2). Improved identification of individuals with AHI, who are most likely to transmit HIV and would have been missed by standard HIV-antibody testing, provides augmented opportunities for same-day ART initiation and behavioural interventions (14). Nonetheless, the study experienced challenges in the uptake and acceptability of the AHI screening and partner notification, as well as the retention in care of the participants diagnosed with AHI –although it is possible that some of these individuals linked to care elsewhere without our knowledge. To improve uptake and retention, we have provided specific training to clinical staff on motivational counselling and we launched a tailored digital community engagement tool (www.cekupyuk.id). Further research is needed to determine the yield of the intervention, mitigate any context-specific implementation barriers, and assess the potential for population impact and cost-effectiveness in the Indonesian context. This information is needed to determine whether AHI screening could be a sensible strategy for wider programmatic implementation. The INTERACT study provides a unique platform to enable future immuno-virological, social science, and intervention studies in Indonesia.

## Data availability statement

The INTERACT Study Group welcome feedback and ideas, including proposals for collaboration on data analyses, new research, or knowledge translation and exchange activities. Requests for data sharing can be made by submission of a study concept to the INTERACT Study Group for evaluation of the scientific value, relevance, design, feasibility, and overlap with existing projects. For more information, please contact the principal investigators: I, KG or RLH.

## Ethics approval

Ethical approval was obtained from the Catholic University of Atma Jaya research ethics committee (0009R/III/PPPE.PM.10.05/10/2022) and the Oxford Tropical Research Ethics Committee (565-22).

## Contributors

I is the principal investigator, and KG and RLH are the co-principal investigators. I, MD, SW, KG and RLH conceptualised the study. I, N, HL, PPJ, FSW, MD, EJS, KG and RLH designed the study protocol. N, HL, S, DR, MO established the cohort and collected the study data and samples. S supervised the laboratory assays. N, HL, MO, DPR managed the clinical database and contributed to data verification. DPR performed the statistical analyses and data visualisations, under supervision of RLH. DPR, RLH and KG drafted the manuscript with critical contributions from I, PPJ, MD, EJS, and FSW. DPR and RLH had full access to all of the study data and take responsibility for the integrity of the data and the accuracy of the data analysis. All authors provided valuable input to interpretation of the data and critically reviewed the paper and figures for important intellectual content. All authors reviewed and approved the final version of the manuscript.

## Acknowledgments

The INTERACT research team would like to acknowledge the contribution of all the participants, the clinic staff at the Globalindo and Bali Peduli clinics, the study support staff at OUCRU Indonesia, and our community partners at Yayasan Inti Muda, Yayasan Gaya Dewata, Yayasan Inter Medika and Yayasan Kasih Suwitno.

**Indonesia Intervention Study to Test & Treat People with Acute HIV Infection (INTERACT) Study Group:**

Prof Irwanto (PI), Ignatius Praptoraharjo, Arie Rahadi, Evi Sukmaningrum (Atma Jaya Catholic University, Jakarta)

Suwarti, Decy Subekti, Bachtiar Andy Mussafa, Nicolas Tarino, Mutia Rahardjani, Fitri Dewi, Soraya Weldina Ragil Dien, Margaret Oktavia, Dwi Rahmawati, Raph Hamers (co-PI)

(Oxford University Clinical Research Unit Indonesia, Jakarta)

Keerti Gedela (co-PI) (56 Dean Street, Chelsea & Westminster Hospital, Imperial College London, UK)

Prof Pande Putu Januraga (Centre for Public Health Innovation, Udayana University, Bali) Prof Evy Yunihastuti (Dept of Internal Medicine, Dr Cipto Mangukusumo Hospital, Faculty of Medicine, Universitas Indonesia, Jakarta)

Nurhayati Agus, F. Stephen Wignall (Yayasan Globalindo, Jakarta)

Hendry Luis, F. Stephen Wignall (Yayasan Bali Peduli, Bali)

Godelieve de Bree, Maartje Dijkstra (Amsterdam UMC, location AMC, University of Amsterdam, Amsterdam, The Netherlands)

Prof Eduard Sanders (The Aurum Institute, Johannesburg, South Africa; and University of Oxford, Sir William Dunn School of Pathology, Oxford, UK)

Sayem Ahmed (University of Glasgow, Glasgow, UK)

Prof Christophe Fraser, Katrina Lythgoe (Big Data Institute, University of Oxford, Oxford, UK)

