## Supplementary materials for "Implementing a clinical pathway for diagnosing and treating acute HIV infection among key populations attending sexual health clinics in Indonesia: cohort profile of the INTERACT study"

**SUPPLEMENTARY MATERIAL**

**Table S1.** STROBE checklist

| **Section/Topic** | Item | Recommendation | Reported on page # |
| --- | --- | --- | --- |
| **Title and abstract** | 1 | (*a*) Indicate the study’s design with a commonly used term in the title or the abstract | 2 |
|  |  | (*b*) Provide in the abstract an informative and balanced summary of what was done and what was found | 2 |
| Introduction | | |  |
| Background/rationale | 2 | Explain the scientific background and rationale for the investigation being reported | 4-6 |
| Objectives | 3 | State specific objectives, including any prespecified hypotheses | 6 |
| Methods | | |  |
| Study design | 4 | Present key elements of study design early in the paper | 7-8 |
| Setting | 5 | Describe the setting, locations, and relevant dates, including periods of recruitment, exposure, follow-up, and data collection | 8 |
| Participants | 6 | (*a*) Give the eligibility criteria, and the sources and methods of selection of participants | 8 |
| Variables | 7 | Clearly define all outcomes, exposures, predictors, potential confounders, and effect modifiers. Give diagnostic criteria, if applicable | 7, Table S4 |
| Data sources/ measurement | 8* | For each variable of interest, give sources of data and details of methods of assessment (measurement). Describe comparability of assessment methods if there is more than one group | 8-10, Table S2 |
| Bias | 9 | Describe any efforts to address potential sources of bias | 8-9 |
| Study size | 10 | Explain how the study size was arrived at | 8 |
| Quantitative variables | 11 | Explain how quantitative variables were handled in the analyses. If applicable, describe which groupings were chosen and why | Table 1, 2, S2 |
| Statistical methods | 12 | (*a*) Describe all statistical methods, including those used to control for confounding | Table 1, 2 |
|  |  | (*b*) Describe any methods used to examine subgroups and interactions | na |
|  |  | (*c*) Explain how missing data were addressed | na |
|  |  | (*d*) If applicable, describe analytical methods taking account of sampling strategy | na |
|  |  | (*e*) Describe any sensitivity analyses | na |
| **Results** |  |  |  |
| Participants | 13* | (a) Report numbers of individuals at each stage of study—eg numbers potentially eligible, examined for eligibility, confirmed eligible, included in the study, completing follow-up, and analysed | 11,  Figure 2,3 |
|  |  | (b) Give reasons for non-participation at each stage | 8-9,  Figure 2 |
|  |  | (c) Consider use of a flow diagram | Figure 2 |
| Descriptive data | 14* | (a) Give characteristics of study participants (eg demographic, clinical, social) and information on exposures and potential confounders | Table 1, Table 2 |
|  |  | (b) Indicate number of participants with missing data for each variable of interest | Table 1, Table 2 |
| Outcome data | 15* | Report numbers of outcome events or summary measures | 11-13 |
| Main results | 16 | (*a*) Give unadjusted estimates and, if applicable, confounder-adjusted estimates and their precision (eg, 95% confidence interval). Make clear which confounders were adjusted for and why they were included | 11-13 |
|  |  | (*b*) Report category boundaries when continuous variables were categorized | na |
|  |  | (*c*) If relevant, consider translating estimates of relative risk into absolute risk for a meaningful time period | na |
| Other analyses | 17 | Report other analyses done—eg analyses of subgroups and interactions, and sensitivity analyses | na |
| Discussion |  |  |  |
| Key results | 18 | Summarise key results with reference to study objectives | 11-13 |
| Limitations | 19 | Discuss limitations of the study, taking into account sources of potential bias or imprecision. Discuss both direction and magnitude of any potential bias | 3,13 |
| Interpretation | 20 | Give a cautious overall interpretation of results considering objectives, limitations, multiplicity of analyses, results from similar studies, and other relevant evidence | 3,13 |
| Generalisability | 21 | Discuss the generalisability (external validity) of the study results | 13 |
| Other information |  |  |  |
| Funding | 22 | Give the source of funding and the role of the funders for the present study and, if applicable, for the original study on which the present article is based | 15 |

**Table S2.** Study outcome measures

| **Study Objectives** | **Main outcome measures** |
| --- | --- |
| **Primary objectives** |  |
| 1. To evaluate the effectiveness/diagnostic yield of an AHI clinical pathway | Descriptive outcomes:   - Proportions of reported symptoms and risk behaviour, and their association with AHI; - Number and proportion of participants who are newly diagnosed with acute, early, recent and chronic HIV, and AHI as proportion of all HIV diagnoses; - AHI and HIV incidence estimations;   Diagnostic yield outcomes:   - Yield of 4^th^ gen HIV/Ag and/or HIV-PCR in diagnosing AHI; - Predictive performance of the Amsterdam AHI risk score, and a localized risk score, expressed as sensitivity, specificity and Area Under the Curve;   AHI screening cascade outcomes:   - Number and proportions of clinic attendees screened for study eligibility, enrolled in the study, screened for AHI, diagnosed with AHI, linked to care, started ART, and achieved viral suppression on ART; - Time (hours/days) from study screening to HIV/AHI diagnosis to ART initiation - HIV-RNA load levels (mean/median and proportion <50 cps/mL) at the time of AHI diagnosis and after 12 and 24 weeks of ART (compared to historical records for all HIV diagnoses); - CD4 T-cell counts (median and categories) at the time of AHI diagnosis and after 12 and 24 weeks of ART   *All outcomes to be compared to historical records for all HIV diagnoses if appropriate and feasible* |
| 2. To evaluate the uptake/adoption, acceptance and feasibility of enhanced HIV testing and partner notification, as part of the AHI clinical pathway | - Number and proportion of individuals who underwent AHI screening, proportion and median repeat AHI screenings, with median time intervals; - Number and proportion of sexual partners of individuals who are newly HIV/AHI diagnosed (index clients), who are notified and if feasible, tested for HIV/AHI and newly HIV/AHI diagnosed; - Barriers and enablers of implementing the AHI clinical pathway from the participant’s and provider’s perspective |
| 3. To estimate the potential population impact and cost-effectiveness of an AHI clinical pathway on curbing the HIV epidemic in Indonesia | - Projections of HIV infections, hospitalizations, and deaths averted and QALYs gained under different intervention scale-up scenarios, including maintenance of the status quo; - Cost-effectiveness of different intervention scale-up scenarios, expressed as ICER measured as cost-per QALY gained |
| **Secondary objective** |  |
| To improve the implementation of the AHI clinical pathway through a tailored digital behavioural/risk assessment intervention and community outreach | - Uptake of a tailored digital risk reduction intervention amongst MSM at risk in the local area in all clinical sites; - Number of digital engagements and length of digital interaction time spent on the tool, and number of returning digital users; - Number of those engaged enrolling in the INTERACT study |
| **Exploratory objectives** |  |
| 1. To estimate the prevalence of transmitted HIV drug resistance in newly HIV diagnosed individuals 2. To understand HIV transmission dynamics in Indonesia | - Proportions of participants with any, reverse-transcriptase (RT), protease (PR) and integrase (IN) resistance, and proportions of major drug resistance mutations; - HIV transmission networks within and between different risk groups and geographic regions, mixing and source attribution, based on phylogenetic inference |

**Table S3.** HIV diagnostic classification

|  | **HIV-PCR** | **HIV-Ag p24** | **HIV-Ab** | **Previous HIV test result** |
| --- | --- | --- | --- | --- |
| **Acute HIV** | + | +/- | - | NA |
| **Early HIV** | + | +/- | Discordant* | NA |
| **Recent HIV** | + | +/- | + | Documented negative test <6 mo prior |
| **Chronic HIV** | + | +/- | + | No documented negative test <6 mo prior |

*Discordant anti-HIV antibodies are defined as: “Combo” Ag/Ab Ab-negative; and third-generation RDT 1x positive and 1x negative OR “Combo” Ag/Ab Ab-positive; and third-generation RDT 2x negative, or 1x positive and 1x negative

Abbreviations: NA, not applicable; RDT, rapid diagnostic test

**Table S4.** Clinical and lab variables

**DEMOGRAPHIC DATA AND ENROLMENT VISIT (S1)**

**Sociodemographic and contact information**

- Data collection date
- Returning or first-time client
- Date of birth
- Nationality
- Sex assigned at birth
- Health insurance
- Marital status
- Education level
- Occupation

**Study eligibility criteria**

Key population/Risk category

- Men who have sex with men
- Transgender person
- Person who injects drugs
- Sex worker
- Client of sex worker
- Having a sexual partner living with HIV
- Other (undisclosed) risk of HIV acquisition

Ever tested positive with HIV

- If yes, specify: currently receiving HIV treatment, and name of the health facility

**Digital consent**

- Do you agree to take part in this study?
- Do you agree to the anonymised use of your medical record and your blood sample, for the purpose of this study?

**Maturity assessment for individuals 16 or 17 years of age**

- It is in the young person's best interest to receive access to HIV prevention and testing services, with or without their parents'/carers' consent
- The young person cannot be persuaded to inform their parents/carers, or allow the practitioner to do so, that they are giving permission to participate in this screening study
- The young person understands the purpose of the study, what is required from participants, the risk and benefits of participating; and is able to make an informed, independent decision about his/her study participation

**FIRST AND SUBSEQUENT AHI SCREEN VISITS (S1, S2, S3, ETC)**

**AHI Risk Checker**

Risk in the past 6 months:

- Number of sexual partners
- STI and or STI symptoms
- Condomless receptive anal intercourse

Symptoms in the past 2 weeks:

- Weight loss (for no known reason)
- Fever
- Swollen glands (lymph nodes)
- Whitish patches (thrush) in your mouth
- Nausea
- Vomiting
- Muscle pain
- Diarrhoea
- Night sweats
- Pain in your joints
- Fatigue/tiredness
- Headache
- A sore throat
- Flu-like symptoms
- Genital warts
- Blisters on your genitals
- Blisters in your mouth

**Sexual behaviour**

- Fixed/regular partner and their gender identity
- Other sex partner (s) and their gender identity
- Date of last sexual intercourse
- Anal sex in the past 3 months;

If yes, specify: condom use and role (insertive/receptive/both)

**Additional questions on the risk of getting HIV**

- Reason(s) for your HIV/STI test today

Feeling at risk

Having symptoms

New sexual relationship

Partner tested HIV positive

Partner tested positive for STI

Retest (window period)

Pregnant or partner pregnant

Getting married

Other

- Current gender identity
- Injected drug use in the past 3 months;

If yes, specify: drugs, needle sharing and frequency

- Recreational drugs before or during sex in the last 3 months

If yes, specify: drugs and frequency

- Sex parties in the past 3 months

If yes, specify: frequency

- Group sex in the past 3 months

If yes, specify: frequency

- PrEP use;

If yes, specify: regimen, access, paid, interest

**Quality of life**

EQ-5D:

- Mobility (scale 1-5)
- Self-care (scale 1-5)
- Usual activities (scale 1-5)
- Pain-discomfort (scale 1-5)
- Anxiety/depression (scale 1-5)
- Health condition today (scale 0-100)

**Counselling and testing**

- Visit date
- History of previous HIV and STI tests
- Current clinician's STI diagnosis
- Current STI treatment provided
- Time HIV and STI results received
- Have clients received post-test counseling?
- If HIV+, specify: linked to HIV care, transfer out, name of health facility

**Lab test result**

- HIV standard tests (R1, R2, R3)
- Xpert HIV-PCR detection assays
- Syphilis RPR/TP Rapid Test and RPR titer
- Xpert CTNG
- HBV HBsAg
- HCV Xpert/Rapid Test

**HIV BASELINE VISIT (T0)**

**Quality of life**

- EQ-5D (see above)

**Assisted partner notification**

- Number of sex partners (including regular partner, casual, one-time encounter or other)
- Assisted partner notification (number of partners to be notified)
- Intimate partner violence assessment
- Partner details (gender identity, age, type of partner/relationship, partner's last HIV test, partner’s HIV status, status of relationship)

**Baseline data**

- Date and time of current visit
- Date and time patient informed of HIV diagnosis
- Enrolled (registered) for HIV care

If other clinic, name of health facility and date of registration

- Pregnancy
- Enrolled in the virologic substudy

**Clinical data**

- Current weight (kg)
- Height (cm)
- HIV symptoms
- Acute retroviral syndrome
- AIDS defining condition or opportunistic infection (if yes, specify)
- TB assessment, diagnosis, treatment
- Antiretroviral use history
- Current ART regimen
- Laboratory data (hemoglobin, creatinine, ALT/SGPT, CD4+ cell count, HIV viral load, other)

**HIV FOLLOW-UP VISITS (T3 AND T6)**

**Quality of life**

- EQ-5D (see above)

**Clinical data**

- Date of previous visit
- Pregnancy
- Current weight (kg)
- AIDS defining condition or opportunistic infection (if yes, specify)
- TB assessment, diagnosis, treatment
- IRIS (Immune Reconstitution Inflammatory Syndrome) in the past 3 months
- Current WHO Performance Stage
- Current ART regimen
- ART missed doses
- Indication for cotrimoxazole
- Other co-medications
- Laboratory data (hemoglobin, creatinine, ALT/SGPT, CD4+ cell count, HIV Viral Load, other)

**Table S5.** Characteristics of the first six participants who were diagnosed with AHI

|  | **#1 (GLJ00074)** | **#2 (GLJ00407)** | **#3 (GLJ00647)** | **#4 (GLJ00649)** | **#5 (GLJ01100)** | **#6 (BPD00062)** |
| --- | --- | --- | --- | --- | --- | --- |
| **Location** | Jakarta | Jakarta | Jakarta | Jakarta | Jakarta | Denpasar (Bali) |
| **Enrolment date** | 26-05-2023 | 30-6-2023 | 28-07-2023 | 28-07-2023 | 16-09-2023 | 29-08-2023 |
| **Demographics** |  |  |  |  |  |  |
| Age (years) | 22 | 29 | 25 | 25 | 27 | 29 |
| Sex at birth | Male | Male | Male | Male | Male | Male |
| Client status | First-time | First-time | Returning | First-time | Returning | Returning |
| Education | Highschool completed | Highschool completed | Highschool completed | University completed | Highschool completed | University completed |
| Occupation | Student | Employed | Employed | Employed | Employed | Employed |
| Key population | MSM | MSM | MSM | Undisclosed | MSM | MSM |
| **AHI risk score** | 2 | 3 | 1 | 2 | 3 | 2 |
| Risk in the past 6 months: |  |  |  |  |  |  |
| ≥3 sexual partners | No | No | No | No | Yes | Yes |
| Condomless receptive anal sex | Yes | Yes | No | No | Yes | Yes |
| STI or STI symptoms | No | No | No | Yes | No | No |
| Symptoms in the past 2 weeks: |  |  |  |  |  |  |
| Fever | Yes | Yes | Yes | No | Yes | No |
| Oral thrush | No | No | No | Yes | No | No |
| Lymph nodes | No | No | No | No | No | No |
| Weight loss | No | Yes | No | No | No | No |
| **HIV testing** |  |  |  |  |  |  |
| HIV Ag/Ab test | Negative | Negative | Negative | Negative | Negative | Inconclusive^a^ |
| Xpert HIV-PCR algorithm | Individual Xpert HIV-PCR | Individual Xpert HIV-PCR | Pooled Xpert HIV-PCR | Pooled Xpert HIV-PCR^b^ | Individual Xpert HIV-PCR | Individual Xpert HIV-PCR |
| Xpert HIV-PCR results | Detected in individual sample (Qual) | Detected in individual sample (Qual) ^c^ | Detected in sample pool and deconvoluted individual sample^c^ | Detected in sample pool and deconvoluted individual sample ^c^ | Detected in individual sample (Qual) | Detected in individual sample (Qual) |
| **PrEP use** | Never | Never | Never | Never | Never | Yes (<1 mo ago, event-driven) |
| **Most recent HIV negative test** | Yes (>6 months ago) | Yes (>6 months ago) | Yes (>6 months ago) | Yes (>6 months ago) | Yes (>6 months ago) | Yes (<6 months ago) |
| **Syphilis** | Positive^d^ | Negative | Negative | Negative | Positive^d^ | Positive^d^ |
| **HIV viral load** | >1x10^7^ cps/mL | ND^c^ | >1x10^7^ cps/mL | 40 cps/mL | >1 x10^7^ cps/mL | >1x10^7^ cps/mL |
| **Assisted Partner Notification** | Declined | ND^c^ | ND^c^ | ND^c^ | Declined | Accepted  (3 partners notified) |
| **Linked to HIV care & started ART** | Yes (same day) | Unknown^c^ | Unknown^c^ | Unknown^c^ | Yes (same day) | Yes (same day) |

^a^ Inconclusive result based on: “Combo” Ag (p24) positive and Ab negative (R1), followed by RDT 2x negative (R2/R3)

^b^ Due to Xpert cartridge stockout, this participant was tested with pooled approach (Xpert HIV Viral Load assay) instead of individual test (Xpert HIV Qual assay)

^c^ Participants did not return to clinic for confirmatory testing and have been lost to follow up. No record could be found in the MOH national HIV/AIDS information system (SIHA).

^d^ Declining RPR titer after recent treatment

Abbreviations: AHI, acute HIV infection; MSM, men who have sex with men; ND, not done; PrEP, pre-exposure prophylaxis; STI, Sexually Transmitted Infection

**Figure S1.** Geographic map of HIV prevalence per province

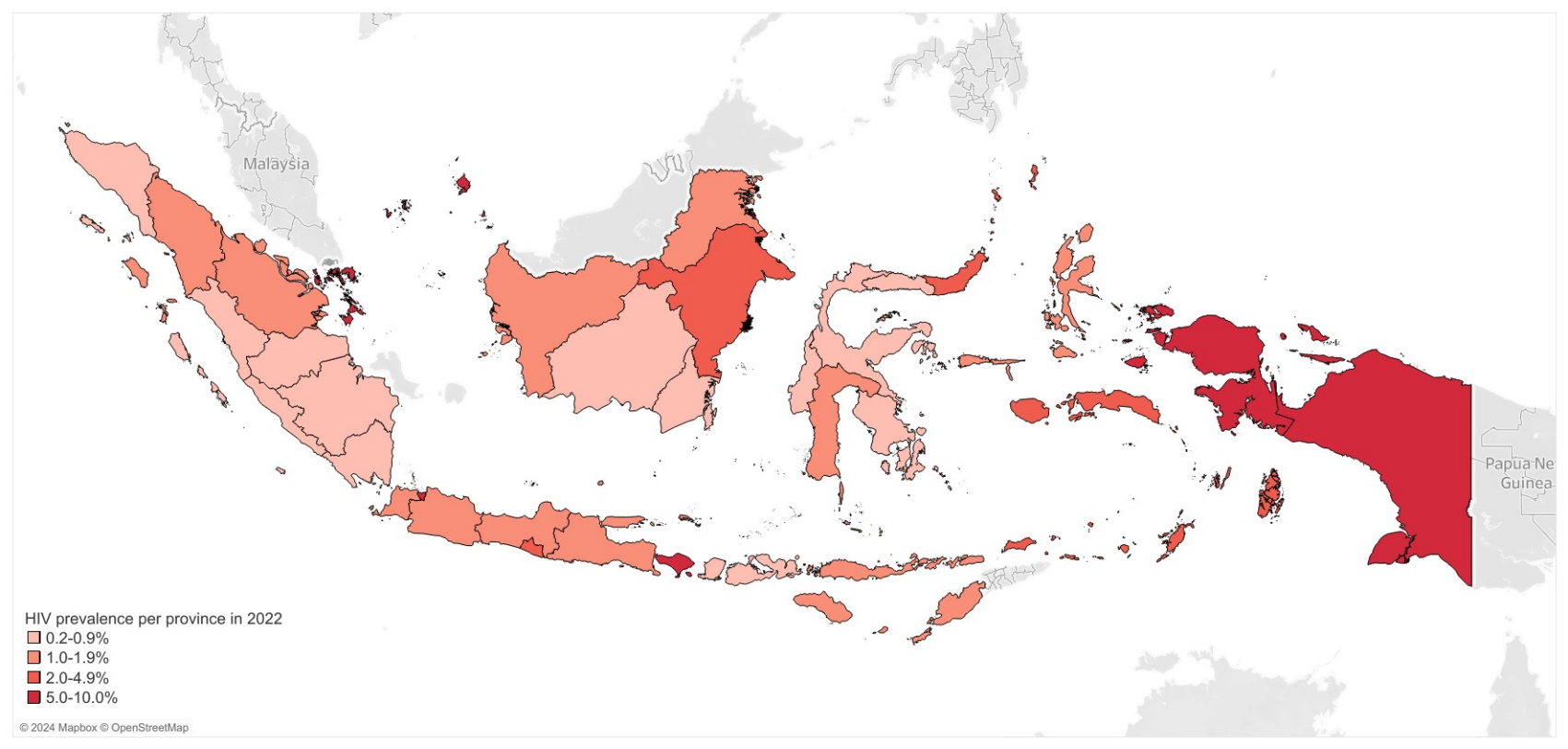

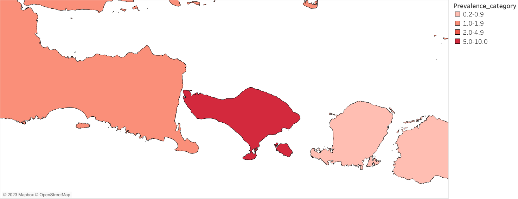

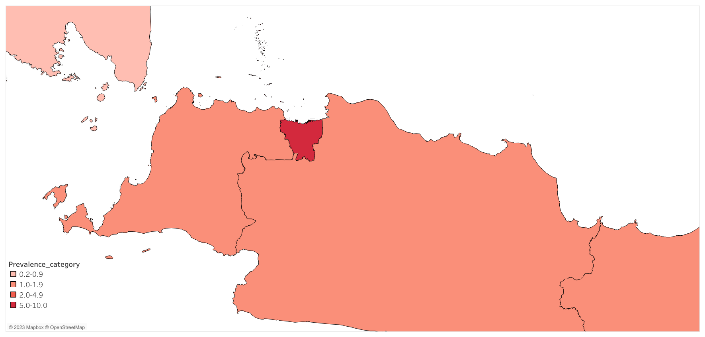

Jakarta

Bali

Figure shows a geographical map of Indonesia with the estimated HIV prevalence for each of the 34 provinces in 2022. The study sites are located in Jakarta and Bali provinces, which have the highest HIV prevalence behind Papua.

Data source: Ministry of Health (2022) (2)

**Figure S2.** AHI test algorithm

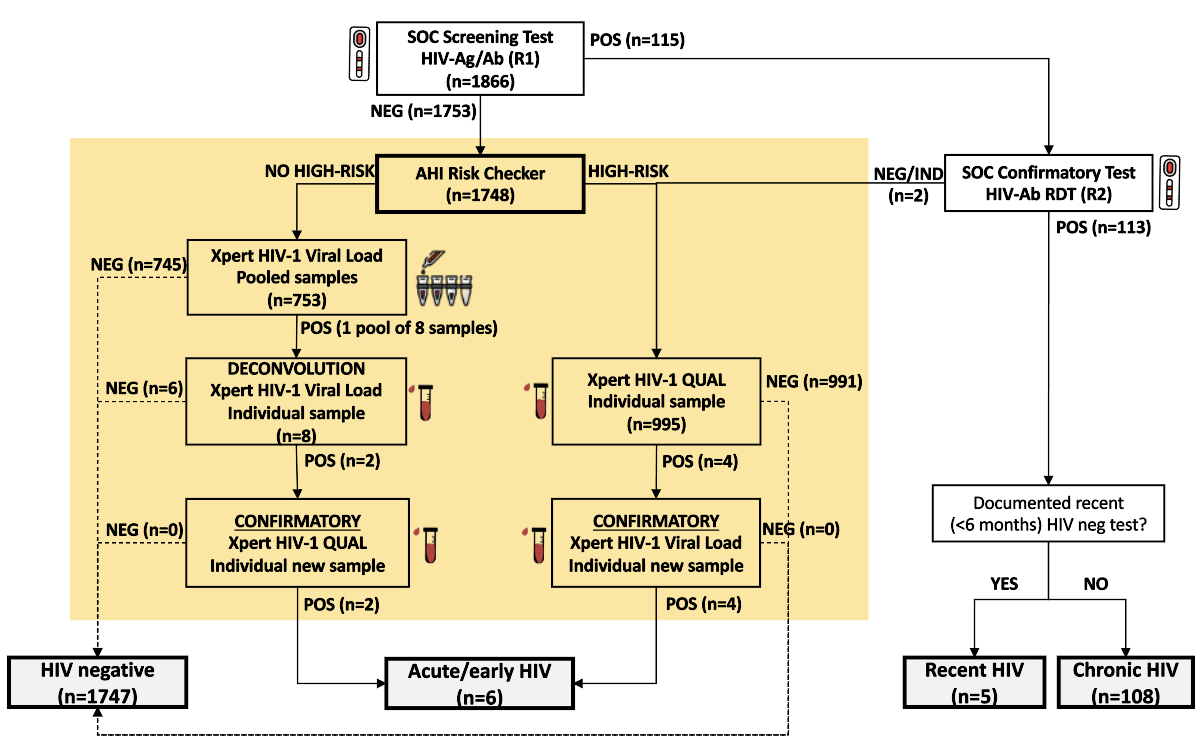

Figure shows the AHI laboratory testing algorithm used in the INTERACT study, including the numbers of samples tested and test results during the first eight months of study enrolment. The section with yellow background represent the AHI screen procedures, comprising the AHI Risk Checker and Xpert HIV-PCR testing. HIV diagnostic classification is summarised in Table S3.

Abbreviations: Ab, HIV antibody; Ag, HIV p24 antigen; IND, indeterminate/inconclusive; NEG, negative test result; POS, positive test result; RDT, rapid diagnostic test; SOC, standard of care;
